# MAIN CAUSES OF MEDICINE STOCK-OUTS IN MAURITANIA: A QUALITATIVE STUDY

**DOI:** 10.1101/2024.05.23.24307794

**Authors:** Mohamed Ali Ag Ahmed, Issa Coulibaly, Raffaella Ravinetto, Verónica Trasancos Buitrago, Catherine Dujardin

**Author notes:** **Corresponding author:** Mohamed Ali AG AHMED. MD, MPH, PhD. **Competing interests** SO. **Authors’ contributions Conceptualization:** MAAA & RR Data curation: MAAA Formal analysis: MAAA & IC Funding acquisition: VT & CD Investigation: MAAA & IC Methodology: MAAA, CD & RR Supervision: MAAA Validation: MAAA, RR, CD Visualization: MAAA & IC Writing – original draft: MAAA, IC, VT, CD & RR Writing – review & editing: MAAA, IC, VT, CD & RR. All authors approved the final manuscript.

## Abstract

The number of medicine stock-outs (MSOs) is increasing globally. In Mauritania, they are recurring, although, to our knowledge, no study has yet been conducted to determine the causes. Therefore, this qualitative study aims to identify the main local or national causes of stock-outs to provide a common understanding and guide policy-makers towards corrective actions. The study was carried out in five health districts and at the regional and central levels. The samples were purposive. Two focus groups and twenty semi-structured individual interviews were held with 38 participants, including health professionals, managers from the Central Purchasing Office for Essential Medicines and Consumables (CAMEC), the Pharmacy and Laboratory Department (DPL) and the Ministry of Health (MoH). All interviews were recorded and transcribed. A thematic content analysis was carried out. Our findings indicate the national causes of MSOs at three healthcare system levels (operational, regional, and central). They were grouped into five categories: insufficient human resource capacity (number of staff, training, retention), communication and coordination problems between stakeholders, logistical constraints (transport, storage), financial constraints, inadequate forecasting of needs, and complex procurement procedures. These causes of MSOs are interconnected, and many could be addressed locally through solutions initiated and led by the Mauritanian authorities. To address MSOs sustainably, we suggest and discuss some possible actions, including reforms to improve CAMEC’s governance and accountability and, more broadly, to strengthen the various pillars of the local health and pharmaceutical system.

## INTRODUCTION

Health is a fundamental human right (1, 2) that cannot be achieved without access to safe and effective essential medicines. The relevance of equitable access to essential medicines has been reinforced by its inclusion in target 3.8 of the Sustainable Development Goals, i.e., Universal Health Coverage (UHC) (3). However, in most countries, and even more so in Africa, access is compromised by increasingly frequent stock-outs of medicines (MSOs) (4, 5, 6). MSOs represent a challenge to providing affordable quality health services and pose risks to patients’ health through nontreatment, undertreatment, and inadequate treatment, including possible medication errors due to attempts to replace missing medicines (6, 7). They undoubtedly contribute to poor health indicators in many African countries, making them a significant obstacle to achieving UHC (6, 7, 8, 9, 10, 11). Identifying the causes of MSOs, which are numerous and complex and may differ by region and country, is essential to prevent and correct MSOs. These problems primarily include production problems and weak supply chains (12, 13, 14); both have been magnified during and after the COVID-19 epidemic, triggering calls in different regions and countries to promote local production, given autonomy and health security. Specific examples include dependence on external suppliers for either active pharmaceutical ingredients or finished pharmaceutical products, poor supply infrastructure, insufficient pharmaceutical budget at the national or local level, political instability, and market dynamic issues (15, 16, 17, 18). More specific causes are long delivery times, delays in awarding tenders, suppliers’ inability to meet demand, or suppliers’ failure to pay (19, 20, 21). Unfortunately, shortages and stock-outs are often magnified during public health emergencies, such as disasters and epidemics. However, they can also be prompted by nonplanned changes in recommended clinical practice.

To strengthen the performance of its pharmaceutical system and—among other things—to minimise the risk of MSOs, Mauritania adopted a national policy and action plan for the pharmaceutical sector in 2022 (22). In this frame, four key actors play an essential role in the supply and distribution of medicines: the Directorate of Pharmacy and Laboratories (DPL); the Central Purchasing Office for Essential Medicines and Consumables (CAMEC - *Centrale d’Achat des Médicaments Essentiels et Consommables)*; private wholesale distributors; and health facilities (22).

The DPL regulates and oversees the pharmaceutical sector and manages all regulatory functions (22). It oversees the implementation of regulatory mechanisms for the marketing authorisation of medicines, the licensing of suppliers, distributors, outlets, and any facilities involved in the supply and provision of medical products. It also draws up the National List of Essential Medicines (NLEM). The current version, issued in 2021, includes 543 medicines. The DPL is also responsible for regulating the import and export of medicines. It oversees public and private sectors, including priority health programs (e.g., AIDS, tuberculosis).

The CAMEC is responsible for making essential quality generic medicines available nationwide. It is the only actor authorised to import some specific medicines for public and private services, namely 39 antibiotics, all insulins, eight oral antidiabetics, 49 cardiovascular products, 11 psychotropic products, seven narcotics, and two thyroid hormones. Private wholesalers must also source these medicines from CAMEC only. The CAMEC has a central warehouse and 12 regional depots from which health facilities obtain their supplies countrywide.

In 2022, 38 private wholesaler-distributors existed in Mauritania, of whom only ten imported health products (22). They supply decentralised ‘sales depots’ and private pharmacies.

Finally, the public health facilities (health centres and health posts) obtain their supplies autonomously directly from the regional depot and, in the case of the health facilities in the capital city Nouakchott, from CAMEC’s central warehouses. However, in the event of an MSO at CAMEC, health facilities can obtain supplies from private wholesalers with special authorisation.

These four key actors face many challenges that limit their ability to play a role in the timely and comprehensive supply and distribution of medicines according to their needs. According to an analysis published in 2020 by the WHO, these challenges include inadequate storage space and logistical resources for drug distribution, the lack of a logistics management information system, and poor quantification of medicine needs (23). Health facilities are also facing specific challenges related to the absence or inadequacy of tools for quantifying and managing stocks (i.e., missing order registers, missing stock sheets or stock sheets not kept up to date, lack of medicine registers), insufficient staff training on stock management and good storage and distribution practices, and a lack of formative supervision (22). To date, MSOs are recurrent throughout the country. According to the 2018 survey “Services Availability and Readiness Assessment” (SARA), the average availability of the 24 WHO standard tracer molecules in Mauritania’s 919 health facilities decreased from 28% in 2016 to 19% in 2018 (24). According to the same survey, only 22% of Mauritania’s public and private health facilities have a stock of 13 tracer medicines (24). Preliminary results from an ongoing study on the availability and prices of medicines in five health districts in Brakna and Nouakchott show similar trends (25). These persistent MSOs in the country underscore the need to explore this issue in more detail. However, to our knowledge, a study has yet to be carried out to determine the causes underlying this phenomenon.

This study is nested in the AI-PASS project “Institutional Support for the Health Sector Support Program”, which was set up in 2017. It is funded by the European Union and implemented by the Belgian Development Agency-Enabel with the support of the Antwerp Institute of Tropical Medicine (26, 27). The project aims to assist the Ministry of Health (MoH) in implementing its National Health Development Plan, covering five components: governance, equitable access to quality care, human resources management, the introduction of a social health insurance scheme, and essential medicines and consumables. Through the last component, “AI-PASS” supports pharmaceutical sector reforms and their implementation at the central level. Moreover, at the operational (decentralised) level and as part of a research-action program (28), it supports the management of pharmacies and the implementation of a medicine supply model within five Moughataa (or health districts). This experience revealed that despite improvements in medicine management and policy action at the national level, stock-outs persisted in several health facilities. In response to these findings, we designed this study to identify the main causes of MSOs in the five health districts where the AI-PASS project is active in Mauritania.

## METHODOLOGY

### Study area and design

Mauritania is a vast desert country covering 1,030,700 km^2^ with a population of 4,649,660 (in 2020) (29). It is one of the least populated countries in the world. It is ranked 164th in the Human Development Index out of 193 countries in 2022 (30). The health system is structured on three levels: the peripheral or operational level, which consists of 57 Moughataa, commonly known as health districts; the intermediate level, which consists of 15 Regional Health Directorates (RHD); and the central level, which consists of the DPL and the CAMEC.

This qualitative study was conducted in five Moughataa, purposively selected in the three health regions where the AI-PASS project is active: Brakna, Nouakchott North and West. Two moughataa (Dar-naim and Sebkha) are located in the country’s capital (Nouakchott), while the other three (Aleg, Bogué and Bababé) are in the Brakna region in the central-southern region of the country, some 300 km away. Each moughataa consists of health posts and health centres that offer a minimum and complementary package of health care and services to the population. The data were collected over one year (November 2022 to October 2023).

### Population, sampling, and recruitment strategy

The study population included health professionals, CAMEC, DPL and MoH managers. Sampling was purposive (31, 32). An initial list of participants and their contacts was drawn up during a workshop held in November 2022 in Nouakchott as part of the AI-PASS project, which united the main actors involved in the supply and distribution of medicines. The inclusion criteria were defined as follows: 1) 18 years of age or older, 2) French speaker, 3) willing and available to participate in the study, 4) involved or significantly experienced in the management or regulation of medicines in Mauritania, and 5) willing to give informed consent. At the start of the data collection phase in November 2022, each participant was called directly by one of the first two authors to request his or her participation. If he or she agreed, an appointment was made to explain the study’s objectives, check whether the prospective participant met the inclusion criteria, and ask permission to participate in the interview. If he or she agreed, the contents of the informed consent form were shared with him or her, and all his or her questions were answered before formal consent was given and the interview began.

Overall, 36 participants were recruited at three levels of the health pyramid—central, regional, and operational—aiming for maximal diversity. Table 1 provides details. We included 12 participants at the central and regional levels and 24 at the operational level. These included doctors, nurses, pharmacists, depot managers, and others.

**Table 1:**
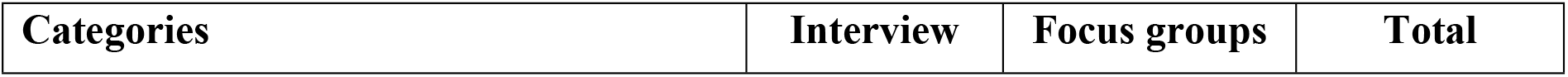
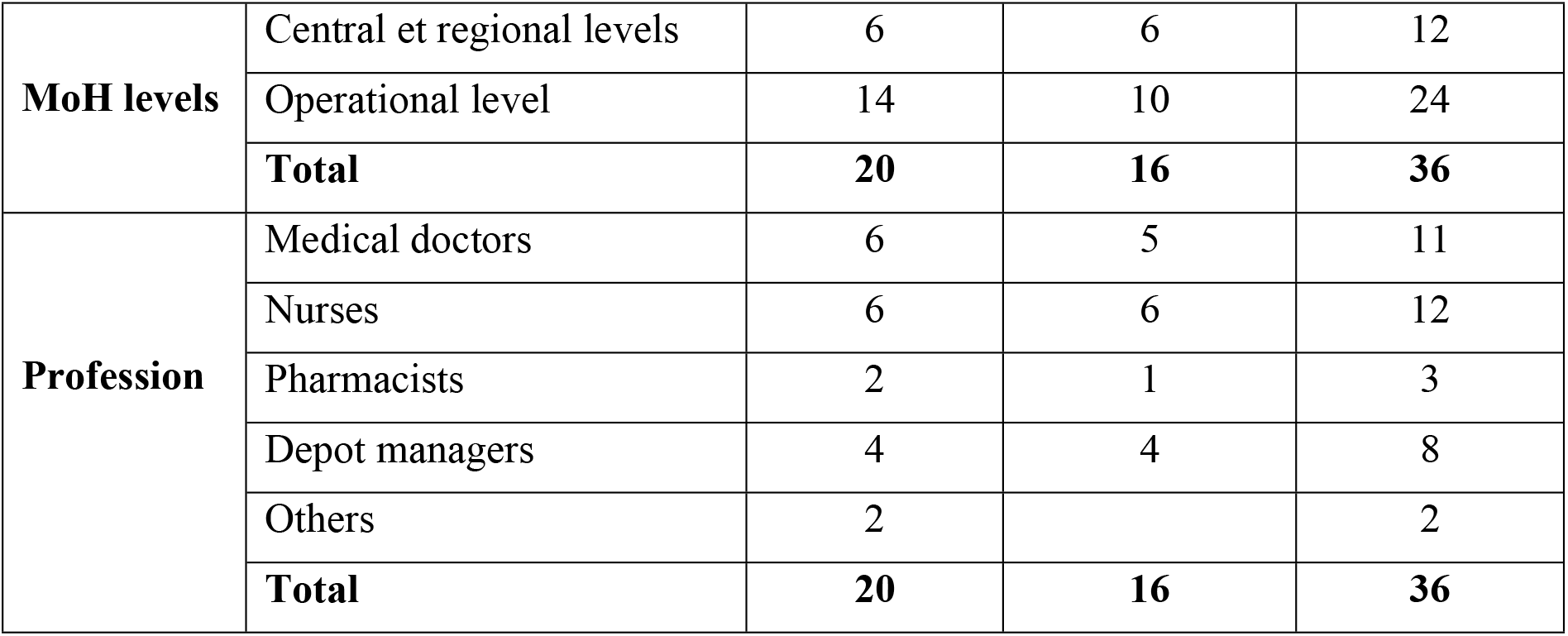
Repartition of participants by MoH levels and profession.

| Categories | Interview | Focus groups | Total |
| --- | --- | --- | --- |

|  |  |  |  |  |
| --- | --- | --- | --- | --- |
| **MoH levels** | Central et regional levels | 6 | 6 | 12 |
|  | Operational level | 14 | 10 | 24 |
|  | **Total** | **20** | **16** | **36** |
| **Profession** | Medical doctors | 6 | 5 | 11 |
|  | Nurses | 6 | 6 | 12 |
|  | Pharmacists | 2 | 1 | 3 |
|  | Depot managers | 4 | 4 | 8 |
|  | Others | 2 |  | 2 |
|  | **Total** | **20** | **16** | **36** |

### Data collection

The first two authors (all PhDs, one medical doctor and the other pharmacist with basic training) conducted two focus groups (eight participants each) and twenty semi-structured individual interviews. These methods are the most widely used data collection techniques in qualitative research and the health care context and enable information to be gathered from key informants with personal experiences, attitudes, perceptions, and beliefs related to the subject of interest (33, 34).

The focus groups were held in conjunction with the November 2022 workshop. Group I consisted of participants from the operational level, and Group II consisted of participants from the regional and central levels. Each group was asked to identify the main causes of MSOs at the operational level for Group I and the regional and central levels for Group II.

The findings from the two focus groups were used to develop the two interview guides for the individual interviews—for the operational level and the regional and central levels. These guidelines included open-ended questions that allowed participants to delve deeper into their experiences of the causes of MSOs. They were pretested on four (non)participants at the peripheral and central levels and refined to capture new emerging issues. The interviews were conducted in French and lasted between 36 and 125 minutes. They were recorded for all participants, as none declined recording during the consent interview. Moreover, notes were taken for each interview.

### Qualitative analysis of the data

Thematic content analysis was used for data analysis (35, 36). The first two authors listened to the audio recordings several times before transcribing them. The second author then conducted a manual analysis using a three-stage inductive approach (coding, thematisation and actual analysis) (36). The verbatims were examined line by line and paragraph by paragraph to generate codes. The closest codes were then grouped under the same subtheme and then under the same induced theme. Ongoing discussions occurred between the first two authors until a consensus was reached on the coding framework. These steps resulted in a thematic tree in which the themes were ranked according to their main or peripheral role. Thematic groupings (convergent, divergent, or complementary) emerged and were presented as meaning matrices that coexist. They enabled us to identify five groups of causes of MSOs in Mauritania, which are reported in our results.

## RESULTS

Participants at the three levels (operational, regional, and central) identified five causes of MSOs: insufficient human resource capacity (number of staff, training, retention), communication and coordination problems between stakeholders, logistical constraints (transport, storage), financial constraints, inadequate forecasting of needs, and complex procedures for procuring medicines.

### Insufficient human resource capacity

Many participants mentioned poor training and retention and insufficient staff in charge of medicine management at the operational, regional, and central levels of the health system as significant contributors to MSOs. Most interviews revealed that the staff in charge of medicine management at the health facility level still needed to improve despite following some training and courses under the AI-PASS project. For instance, several heads of health posts and some pharmacy depot managers cannot use management tools correctly and do not know how to calculate their average monthly consumption (AMC), resulting in medicine orders that do not match actual needs. In addition, because of insufficient training, orders are not placed on time, contributing to MSOs. A participant from the central level explained it well:

> *“At the* health facilities *level, it is difficult for the heads of post to estimate needs. They do not have the tools they need to determine the molecules’ AMC based on which orders are placed. And not all pharmacy depot managers are trained in medicines management”*.

In the country’s interior, little retention of staff who have received training in medicine management within health facilities was noted. Most of these staff members are transferred to the capital after a short time, and the new staff members, who need to be sufficiently trained to manage medicines, make erroneous estimates. In addition, due to various factors, including poor motivation, lack of integrated supervision (or specific supervision for medicines) and a heavy workload, vigilance regarding monitoring pharmaceutical stocks is decreased. This reduction negatively affects the essential task of keeping stock records current. As a participant from a health centre pointed out,

> “*We only have one person in the pharmacy. Imagine if she had to take care of sales and keep stock records. It is much work for one person, especially with the number of patients we see here daily - on average, 100. There are days when she does not fill in the stock cards*”.

Some participants also mentioned a need for more knowledge of the therapeutic indications of certain medicines (sometimes due to the use of different commercial names) and the failure of poorly trained staff to match the orders with therapeutic protocols. This shortcoming leads them not to order certain molecules despite the product’s availability at CAMEC. As a participant from a health post said,

> *“There is a lack of knowledge of the product; someone who does not know the indication of a product will not order it. Although the product is available from CAMEC, perhaps the product’s name is in a scientific denomination or other names; for example, I am used to working with Buscopan rather than Spasfon. Even if CAMEC has Spasfon, I am not used to it because I am not familiar with that product. So, I am not going to order*…*”*.

According to many participants, CAMEC also needs more qualified staff at the regional and central levels. One participant told us that CAMEC’s regional office comprises a manager, an office assistant, and a caretaker. None of these staff members are pharmacists – as all CAMEC’s pharmacists are in Nouakchott. The manager does not have a delegation line in case of absence due to illness or annual leave. This situation leads to errors in the planning and coordination of supplies, thus contributing to stock-outs of medicines. As a participant confided to us:

> *“CAMEC lacks qualified staff to manage supply flows effectively, which leads to frequent stock-outs of medicines*.*”*

In addition, the need for qualified staff makes accurately estimating the needs of essential medicines impossible. It leads to delays in processing orders, as a participant from a health centre noted:

> “*Delays in processing orders at CAMEC are frequent because of the lack of qualified staff, leading to undesirable stock-outs*”.

### Poor communication and coordination

The participants revealed that due to ineffective communication between the concerned stakeholders, health facilities are often not aware of the medicines available at the central level, which results in inefficiency and wasted time, as described by a participant from a health post:

> *“We don’t know what is available at CAMEC. The list of medicines at CAMEC is not shared. It’s important to know this list so that we can place orders. If we know some products are not available, others can replace them. It’s easier*.*”*

To remedy this situation, some participants welcomed the recent creation of a WhatsApp group by a CAMEC regional representative. In this group, they exchanged information on new stock entries, interactions between certain medicines, products that were close to their expiration date, and the possibility of stock transfers between health facilities. However, this group is informal and restricted because not all health facilities have access.

Participants also pointed to difficulties in effectively communicating their medicine’s needs due to gaps in existing communication channels, as explained by a participant from a health post:

> *“We find it difficult to communicate our essential drug needs effectively”*.

Poor communication among stakeholders results in inaccuracies and delays in the transmission of medicine needs, leading to stock-outs, as mentioned by a participant from the regional level:

> *“At the health centre level, it’s mainly the administrative procedure for supplying medicines to CAMEC. You see, the health centre has to issue its medicine order form a week before proceeding with the other aspects of delivery. The order can go for a month without being fulfilled, even here in Nouakchott. What can I say about what happens in the rest of the country? So this is a problem for CAMEC because it has yet to open regional branches in Nouakchott so far, unlike what it has done in the interior. In Nouakchott, we get our supplies from all the other regions at the central level, including health posts. That is a big problem*.”

Participants also pointed to a lack of coordination between the health facilities and the central purchasing office at CAMEC, leading to gaps in management and unforeseen shortages of medicines; as a participant at the regional level noted:

*“Coordination between levels is a challenge. Information on real needs does not circulate quickly enough, which leads to difficulties in planning supplies”*.

Participants also noted a need for coordination within CAMEC itself. They mentioned that the regional CAMEC has no information on the stock of medicines available at the central level or their quality, and it is not notified of shortages or withdrawals of medicines from the market. For these respondents, the interconnection between the CAMEC central and regional levels is lacking except for the notification of receipts, which occurs via a Dropbox account.

Significant communication and coordination problems at the central level were also mentioned, particularly between CAMEC and key stakeholders such as the regional health directorates and the DPL, including for the supply and distribution of medicines for the vertical programs. Some respondents described uncooperative relations that hindered the flow of information and processes linked to the distribution of medicines, as this respondent from the central level noted:

> *“There is no feedback from CAMEC to the DPL or the* RHD. *There is a total lack of coordination between the various parties involved in supply. Tasks are not clearly defined between those involved. The system has not been working properly for nearly five years. We need to re-examine the role of each organisation”*.

### Major logistical constraints

The participants mentioned that CAMEC may fail to deliver medicines to health facilities due to significant logistical challenges. Enormous difficulties exist in transporting medicines over long distances within the country. Medicines are often transported via private vehicles, ambulances, or unsuitable public transport, which can affect their quality due to exposure to high temperatures, direct sunlight, and dust. This testimony from a participant at a health post is quite illustrative:

> *“I have a friend who comes from Makhta Lahjar. I have to go with him because I know that if I do not go with him, it can take me a long time to get back. So, I have to call him at night. He’s going to book me a place there and back. So, he will take me to CAMEC; I’ll pick up my products, and he’ll bring me back. The transport conditions are also terrible. That’s why I told you that even if the product is of good quality, it is at risk if it’s exposed to the sun for an hour”*.

Several participants believe most health facilities need a dedicated and suitable area for storing medicines. Medicines are often stored in consultation rooms, which must comply with good storage practices. In addition, the health facilities that do not have electricity are forced not to order and store heat-sensitive medicines, as a participant from a health post told us:

> *“There are also concerns about storage conditions. Even in Nouakchott, it’s 45 degrees. There are many posts where my colleagues don’t have air conditioning, and there are many things, so we cannot order a lot when we don’t even have storage space. Plus, the temperature is high, and the building isn’t suitable for storing such large quantities”*.

Participants at the central level also raised this problem of the availability and appropriateness of storage areas, who mentioned CAMEC’s limited capacity to store enough medicines to meet the country’s needs. Additionally, CAMEC does not have a generator to ensure the conservation of products in the event of a power cut. At the regional level, a warehouse cannot hold more than an estimated two-month stock. This situation has forced its manager to order only a small quantity of each medicine, resulting in recurrent stock-outs.

The interviews also revealed that good storage practices are not followed because CAMEC’s infrastructure could be more suitable. For example, one participant told us that CAMEC’s regional office inherited the premises of the former regional hospital, which lacked a false ceiling under the roof. The shelves are not aligned with the windows to prevent the sun’s rays from affecting the products. Moreover, the air conditioning is not working correctly. A participant from the regional level said:

> *“The shop is too small, on average 150 m3, and we were told that action was being taken, but still nothing. The standards are not being met. There is no air conditioning or ventilation in the store. “*.

Products are distributed from the CAMEC central to the regional level using the CAMEC’s refrigerated vehicles. However, CAMEC needs more vehicles (four lorries should serve the entire country) to ensure that medicines are regularly transported from Nouakchott to the regional offices.

### Financial constraints

Many participants stated that health facilities have had problems ensuring proper supply for more than a decade due to inadequate working capital. Without adequate financial resources, their supply capacity remains limited, as a participant from a health centre noted:

> *“These working capital funds were designed around ten years ago and no longer cover the needs of a monthly payment for medicines, as the price of medicines has risen considerably*.*”*

On the other hand, some participants noted that the AI-PASS project and the Ministry of Health, through *Mouyassar*, have established medicine allocations that have helped to recapitalise the working capital. However, some also noted that some delivered medicines may fail to match the needs of health facilities. For example, the facilities received medicines that the health care staff could not prescribe, which led to their expiry on the shelves.

Our interviews revealed that health facilities face the problem of debt recovery. This problem is linked to the introduction of the “obstetrical package” to the public system a few years ago, which was based on prefinancing the cost of pregnancy and childbirth care through a lump sum payment. This system, despite the urgency of providing access to pregnancy and childbirth care, has not been balanced by other financial measures and thus significantly contributed to the decapitalisation of health facilities’ drug sales depots. Some health facilities are two or even three years behind in their payments, as a participant from a health post told us:

> *“We have a theoretical working capital because everything is swallowed up in the obstetrical package. Pregnant women take almost half our medicines, and we must wait months and months for reimbursement. However, they don’t give us credit to buy the medicines, you see*…*”*.

At the CAMEC level, participants also described difficulties in maintaining adequate levels of supply due to limited financial resources, as noted by a participant from the central level:

> *“We face a chronic shortage of funds which hampers our ability to purchase sufficient drugs”*.

Furthermore, CAMEC does not participate in group purchasing mechanisms with other countries, although proposals have been made to move in this direction. These mechanisms could allow CAMEC to negotiate better prices (or purchasing conditions) from manufacturers and other suppliers and prevent fluctuations in medicine prices.

Delays in the payment of suppliers by CAMEC were also highlighted, which negatively impacts CAMEC’s relationship with its suppliers, as mentioned by a participant from the central level:

> *“Late payments have become commonplace, complicating our relations with suppliers and hampering our supplies”*.

Other participants confirmed that CAMEC’s financial difficulties have made negotiations more difficult, compromising the organisation’s credibility towards external counterparts. However, some interviews reported that with the support of the State and the payment-in-advance mechanism in place, CAMEC now has fewer financial resource problems than a few years ago. Moreover, CAMEC no longer sells on credit, which, according to some respondents, has significantly improved CAMEC’s working capital.

### Inadequate forecasting of real needs and complex procedures for procuring medicines

According to some participants, regional CAMEC offices must fully grasp health facilities’ real needs. They noted that CAMEC needs to consult the health facilities or the chief doctors of the moughataa to discuss their actual consumption or needs before procuring supplies. As a result, needs are expressed outside the actual needs of the health facilities. Moreover, the regional CAMEC offices tend to issue orders based on availability data from the regional level only, without consulting at the central level. Consequently, health facilities may experience stock-out of products that are not available at the regional level but are in stock at the central level, as noted by a participant from a health centre:

> *“We sometimes order products unavailable at the regional level. But when I go to Nouakchott for workshops and visit the central CAMEC, I’m told that the products in question are available”*.

For the central CAMEC, participants told us that it consults little or not at all with its regional branches and the DPL about actual needs. As a result, frequent incorrect estimates result in medicine shortages. A participant at the national level mentioned the following:

> *“The national quantification commission is not functional, and health facilities data does not go back to CAMEC. As a result, CAMEC orders quantities that are not quite right*…*”*.

Furthermore, CAMEC also appears to need help making the products available to health facilities and private wholesalers under its monopoly. Some respondents suggested that CAMEC prioritises private wholesalers with considerable financial resources to the detriment of public health facilities, which provide care to a large part of the population. Some even recommended abolishing this monopoly, even if only for medicines for chronic pathologies, and refocusing CAMEC’s missions on the availability of other products.

Moreover, CAMEC procedures are complicated, leading to significant delays in the distribution of medicines, as mentioned by a participant at the regional level:

> *“The administrative steps are so complex that they considerably slow down our ability to obtain the necessary medicines quickly”*.

The delays generated by administrative procedures, with several levels of approval, were mentioned by participants as significant obstacles to procurement and drivers of stock-outs. The medicine-purchasing procedure is based on restricted calls for tender, and all CAMEC medicine purchases are subject to approval by the Market Commission. These measures are essential to protect the community from substandard and falsified medicines. Nevertheless, unpredictable delays may occur if the system performance could be better (for instance, due to a lack of sufficient resources). As a participant from a health centre observed,

> *“Each bureaucratic step adds time, and this extra time often results in stock-outs*.*”*

## DISCUSSION

This study is the first conducted in Mauritania to describe the national causes of medicinal stock-outs at the central and peripheral levels. We explored the views and perspectives of health care and other professionals involved in regulating, supplying, and distributing medicines. Our findings provide in-depth insight into the causes of stock-outs at three levels of the public health system (operational, regional and central). They will inform policy-makers and donors in Mauritania about the necessary corrective measures.

The causes of stockouts are multifactorial and interconnected. Based on thematic analysis, we grouped these causes into five categories: insufficient human resource capacity (number of staff, training, retention); communication and coordination problems among stakeholders; logistical constraints, i.e., at the level of transport and storage infrastructure; financial constraints; and inadequate forecasting of needs coupled to complex procurement procedures.

Qualitative studies of the national causes of MSOs of this type are essential if we devise solutions that are more likely to be found locally (37). However, MSOs are a recurring problem in many countries, especially those in sub-Saharan Africa (SSA), despite increased investment in the purchase of medicines and the introduction of measures to encourage the local production of generic medicines on the continent (37, 38, 39). Mauritania does not manufacture medicines and is consequently 100% dependent on imported health products. For most African countries, pharmaceutical imports represent up to 70-90% of medicines consumed, which proves their high level of dependence and predisposes their populations to drug shortages (40). However, Mauritania’s regulatory system still needs to mature, and the country seems to need more time to invest in local production. The stock-out problem could be addressed by acting at other levels of the pharmaceutical and health system.

First, this study highlights the insufficient human resource capacity at health facilities and CAMEC. This finding is in line with the results of the health system mapping conducted in 2020, which showed that only 65.7% of the positions at health centres and posts were filled by state-qualified nurses, and only 49% were filled by medical social nurses, who are generally responsible for managing drug depots (41). The mapping also highlighted the uneven distribution of health care staff, with approximately 1/3 of health human resources located in the capital, to the detriment of the regions, and the lack of training for pharmacists and pharmacy technicians (the few pharmacists have been trained abroad and remain too few to meet the country’s needs). These human resource issues are a real challenge for Mauritania’s health care systems and most sub-Saharan African countries (42, 43, 44) and hamper the availability of medicines (45, 46). Indeed, other authors have shown a link between poor availability of medicines and low staffing levels or poor qualifications (45, 46, 47) and argued that available, well-trained human resources are a driving force behind supply chain performance and the prevention of stock-outs of essential medicines (45, 48, 49, 50), at least when the causes of stock-outs are located in a country. While the challenges related to staff availability and retention are well known in Mauritania, most participants within the five districts supported by the AI-PASS project surprisingly perceived and insisted on the need for training in pharmaceutical management. The AI-PASS project deploys considerable resources to offer several training courses on medicine management. It has established local mentoring by technical assistants in each of the five health districts to strengthen the day-to-day capabilities of health staff. The lesson learned here is that training alone is not sufficient if trained staff are not part of a well-oiled and not-overloaded team with adequate communication and connection to peers at other health system levels.

Indeed, problems of communication and coordination among stakeholders, as well as transparency on the distribution and supply of medicines, have been highlighted by most participants as significant causes of stockouts. More broadly, these findings raise the question of interprofessional collaboration (51, 52), essential for ensuring quality care for the population, including an efficient medicine supply and distribution system. Such problems are recurrent in SSA, probably due to a weak culture of written communication, a lack of adequate means of communication, and sometimes rigid hierarchical links that are not conducive to good communication and coordination. In the case of pharmaceutical stock management, the lack of communication and coordination results in inaccurate estimates of stocks and needs and inadequate orders and stock planning (46).

Third, our findings show that the logistic weakness of the distribution chain is a significant contributor to MSOs in Mauritania. Most participants indicated that CAMEC’s available infrastructure needs to be improved to cover a territory as vast as Mauritania. It does not enable it to make essential medicines available to the last mile. Moreover, CAMEC supplies medicines only up to the regional level, which prevents or remediates shortages and MSOs in peripheral health facilities. Other studies conducted in Africa similarly indicated that weakness in the pharmaceutical distribution chain is at least partly caused by insufficient, uncoordinated, underfunded and non-standardised logistics and poor storage and distribution practices and appears to be one of the main causes of MSOs (53, 54, 55, 56).

Fourth, in Mauritania, MSOs result from insufficient and inadequate infrastructure and financial resources at the level of both health facilities and CAMEC. Such problems are recurrent in many African countries (38, 46). In Mauritania, insufficient storage areas and inadequate storage conditions at the peripheral, regional and central levels hamper ordering sufficient quantities of medicines. In addition, financial constraints at both health facilities and CAMEC mean that these institutions need more working capital to order medicines regularly and in line with actual needs. In many SSA countries, these challenges are present and exacerbated by low public investment in the pharmaceutical sector and health insurance coverage (57, 58).

Finally, the poor performance of medicine procurement procedures, the lack of willingness or commitment to join regional initiatives for group purchasing, and the inadequate estimates of actual needs found in our study have also been documented as causes of stockouts in other African contexts.

Overall, this study shows that the in-country causes of MSOs are diverse and interconnected. Moreover, it raises questions about the performance of the model of a public purchasing centre, such as CAMEC, and of a central pharmaceutical management and distribution system, which is certainly justified but needs substantial improvement. Unfortunately, the private sector needs to be better regulated, and it tends to serve its interests. Moreover, once these public models have been established, they become so powerful that they are difficult to control or even reform, which can be problematic (38, 54, 56). Long-term needs include transformative changes to go beyond traditional rigid schemes and build a flexible and well-regulated system that can fulfil its public health role by delivering medicines when and where needed.

However, when examining the specific problems of MSOs in Mauritania, practical solutions must be found urgently. CAMEC must define a clearer vision and an accountability mechanism to fulfil its health care mandate. Similarly, the development of a National Drug Supply and Distribution Strategy for Mauritania remains a priority, as does the allocation of substantial resources for its implementation.

At the same time, increasing focus should be placed on strengthening the various pillars of the health care system (59, 60), which needs high-quality initial and in-service training (e.g., through integrated supervision) and production by schools and faculties of sufficient numbers of health care personnel, including pharmacy technicians and pharmacists, to meet the country’s coverage needs. In addition, we need to ensure the equitable distribution of medicines throughout the country by implementing more rigorous, decentralised management and incentives to improve their retention. Substantial investment in infrastructure (construction, renovation, and equipment) will also be essential to ensure that medicines are stored adequately at health facilities or CAMEC. Effective coordination and pooling of the MoH’s efforts with its technical and financial partners can help to implement these solutions.

Establishing transit depots at the moughataa level could be a short-term alternative for bringing medicines closer to health facilities until the technical conditions are met for creating sales depots within the moughataa, as in most neighbouring countries (61). The need for sales depots at the moughtataa level is more acute in regions other than Nouakchott, given its proximity to the central CAMEC. Capacity-building for the management teams running the moughataas’ activities could make a lasting contribution to this objective. Investments are also needed to gradually switch from the current passive mode of transport to an active mode so that medicines can be transported to health facilities or, at the very least, to the health centres that head up the moughataas. Additionally, the digitisation of a management and communication system and the development of a logistics information system capable of providing complete data from all levels of the supply chain (central, regional, Moughataa, and health facilities) should help to rationalise the supply chain and facilitate efficient and transparent communication between all actors (61, 62).

In most SSA countries, most patients pay for their medicines out of pocket, while less than 20% (17.8%) of the population is covered by at least one social protection benefit (63). In this sense, the recent creation of the National Solidarity and Health Fund “*Caisse nationale de solidarité et sante* (CNASS)” in Mauritania, which aims to cover more than 70% of the population in the informal sector, is a source of hope (64). An agreement between the CNASS and CAMEC has been signed to help improve the supply of medicines to member health facilities. In addition, the CNASS should help guarantee more stable resources for health facilities, thus preventing their decapitalisation. These non-exhaustive courses of action could be implemented as part of the ongoing supply chain review to improve the last-mile medicine distribution (61). Finally, new studies are needed to assess the availability and prices of medicines, the causes of shortages of program medicines, and, later, the impact of current reforms and the CNASS on the availability of medicines in health facilities.

## LIMITATIONS

This study does not address the well-documented international causes of drug shortages (15, 65, 66). It focused on national causes that can be addressed locally by the country’s authorities. It explored the points of view of those involved in the national supply and distribution of medicines. However, the views of patients and technical and financial partners were not solicited, which could have provided a different perspective on the causes of MSOs. Additionally, the study focused on the five moughataa where the AI-PASS program operates, and our results consequently cannot be generalised to the entire country, let alone to other SSA countries. However, because of several contextual similarities, lessons can be drawn for Mauritania and other SSA countries. The study could be extended to gather stakeholders’ views on the solutions to remedy this situation. Finally, we are aware that the position of external researchers in this study and the inclusion of the partner Enabel may have influenced the data collection and analysis. To reduce potential bias in the selection of interviewees, we adopted a reflexive and iterative approach, with systematic and rigorous documentation of the data collection and analysis process. Data triangulation and the use of different sources mitigated the risk of bias.

## CONCLUSION

This qualitative study provided evidence on the national (central and peripheral) causes of stockouts within five purposively selected health districts in Mauritania. The identified causes are diverse but strictly interconnected: insufficient human resource capacity, poor communication and coordination between stakeholders, logistical and financial constraints, and inadequate or poorly performing procedures for medicine procurement, supply and management. It also allowed us to identify a series of corrective actions, which should be initiated and led by the Mauritanian authorities, with the support of external donors and actors. Furthermore, focused operational research will be essential to monitor and evaluate these courses of action.

## Data Availability

The raw data from this study are interview transcripts, which contain potentially identifiable and sensitive information about participants. We are unable to share this data publicly due to restrictions imposed by the Institutional Review Board of the Antwerp Institute of Tropical Medicine, as participants have not consented to their data being shared outside the study team. Relevant de-identified extracts from the transcripts are included in the article. Requests for further information can be sent to the Institutional Review Board of the Antwerp Institute of Tropical Medicine at.

## Acknowledgements

We would like to thank the Mauritanian health authorities and the technical assistants (Dr Andrianjafy Tahina Simon et Dr Sophie Jacqueline, Dr Amadou Kane, Dr Mohamed El Oumrani and Cheikh Tourad Abderrahmane) of the AI-PASS project, who efficiently helped to facilitate this data collection.

## Abbreviations

AMC: average monthly consumption
AI-PASS: project Institutional Support for the Health Sector Support Program
CAMEC: (*Centrale d’Achat des Médicaments Essentiels et Consommables):* Central Purchasing Office for Essential Medicines and Consumables
DPL: Pharmacy and Laboratory Department
MoH: Ministry of Health
MSOs: medicine stock-outs
NEML: national Essential Medicines List
HDI: Human Development Index
INRUD: International Network for Rational Use of Drugs
RHD: Regional Health Directorates
RLSs: resource-limited settings
SARA: Services, Availability and Readiness Assessment
UHC: Universal Health Coverage
WHO: World Health Organization

## Declarations

### Ethics approval and consent to participate

The project was carried out by the principles set out in the World Medical Association’s Declaration of Helsinki: ethical principles for medical research involving human subjects (2013). It received ethical approval from the Institutional Review Board of the Institute of Tropical Medicine of Antwerp in Belgium (Ref. No. 1640/22). In the absence of an ethical committee in Mauritania, approval was obtained from the Ministry of Health of Mauritania (Ref N ° 000208/2022). All participants gave their approval and signed the informed consent.

### Consent for publication

SO

### Availability of data and materials

The datasets used and/or analysed during the current study are available from the corresponding author on reasonable request.

### Funding

The European Union financed this study in Mauritania through the AI-PASS project implemented by Belgian Development Agency-Enabel.

## Notes

### Competing Interest Statement

The authors have declared no competing interest.

### Funding Statement

Yes

### Author Declarations

The project received ethical approval from the Institutional Review Board of the Institute of Tropical Medicine of Antwerp in Belgium (Ref. No. 1640/22). In the absence of an ethical committee in Mauritania, approval was obtained from the Ministry of Health of Mauritania (Ref N ° 000208/2022). All participants gave their approval and signed the informed consent.

